# Intoxication-Related Deaths at a Poisoning Referral Center in Isfahan, 2019–2023: Demographic and Other Factors

**DOI:** 10.1101/2025.08.14.25333683

**Authors:** Nahid Yousefian, Shiva Samsamshariat, Gholamali Dorooshi, Nastaran Eizadi-Mood

**Affiliations:** Department of Clinical Toxicology, School of Medicine, Clinical Toxicology Research Center, Isfahan University of Medical Sciences, Isfahan, Iran

**Keywords:** Morbidity, Mortality, Poisoning, Substance-Related Disorders, Suicide, Methadone

## Abstract

**Background:** Acute poisoning is a major public health concern and a significant cause of hospital mortality worldwide. Poisoning-related mortality is influenced by demographic and socioeconomic factors, with suicide being the predominant cause in many regions. This study examines poisoning-related deaths in relation to demographic and other relevant factors over a five-year period in Isfahan, Iran.

**Material and Methods:** A retrospective cross-sectional study was conducted on acute poisoning-related deaths recorded at the Clinical Toxicology Department, the referral poisoning center at Khorshid Hospital, Isfahan, Iran, from March 20, 2019, to March 20, 2023. Data were collected on demographic characteristics, cause of poisoning, type of substance, route of exposure, duration of hospitalization, history of suicide attempts, and addiction. Statistical analyses were performed to assess associations between demographic variables and poisoning outcomes.

**Results:** Among the 306 poisoning-related deaths, 81.4% were males with a median age of 42 years. Suicide accounted for 63.07% of cases, followed by accidental poisoning (18.95%) and substance abuse (17.97%). The most common substances involved were methadone (20.56%), aluminum phosphide (18.63%), and paraquat (17.32%). Ingestion was the primary route of exposure (94.44%). Significant associations were found between gender and route of exposure (*P*=0.043) as well as substance abuse history (*P*=0.024).

**Conclusion:** Suicide was the leading cause of poisoning-related death. Mortality is observed more in patients with methadone, aluminum phosphide, and paraquat poisoning. Preventive measures, including stricter regulation of pesticide sales, monitoring of addiction treatment programs, and enhanced mental health support, are essential to reduce mortality and morbidity from acute poisoning.

## Introduction

Poisoning is a major public health concern worldwide and represents one of the leading causes of hospital emergencies in many countries (Albano et al., 2022). It can lead to both morbidity and mortality, with outcomes influenced by factors such as age, gender, socioeconomic status, geographic location, and the type of substance involved. Patterns of poisoning also vary across regions and demographic groups (Mew et al., 2017).

Research indicates that males are at a higher risk of fatal poisoning in most parts of the world (Slavova et al., 2015). In Karachi, Pakistan, young adults are disproportionately affected by poisoning fatalities (Amir et al., 2019), whereas in developed countries, the affected age groups vary. For example, a study in the United Kingdom reported a mean age of 40–49 years among poisoning deaths (Camidge et al., 2003). In Iran, unintentional poisoning mortality rates were higher due to limited access to healthcare resources and the widespread presence of hazardous substances in some regions (Rasouli et al., 2011). Additionally, pesticide poisoning, whether through misuse or self-poisoning, is a significant concern in these regions (Mew et al., 2017).

Poisoning is a major cause of hospitalization in Iran, with opioids being the most common cause of fatal poisoning (Moradi et al., 2016). Factors associated with the risk of fatal poisoning include age, gender, suicidal intent, unintentional poisoning, socioeconomic status, and regional variation (Mehrpour et al., 2018). Most poisoning cases occur in young adults, with intentional poisoning being the predominant cause. However, in the elderly population, poisoning is more often accidental and complicated by comorbidities (Azizpour et al., 2016). Fatal poisonings are more frequent in men, whereas women are more commonly involved in intentional poisoning cases (Hassanian-Moghaddam et al., 2014). Low income is also a significant risk factor, as it limits access to healthcare and contributes to unsafe storage of toxic substances (Babakhanian et al., 2020). Additionally, rural areas report higher incidences of pesticide poisoning, whereas urban centers such as Tehran and Isfahan exhibit higher rates of opioid-and drug-related fatalities (Titidezh et al., 2019).

Given the critical importance of saving lives in cases of acute poisoning, we conducted a five-year cross-sectional study to investigate the frequency of poisoning-related mortality. We further examined the epidemiological and intoxication-related factors influencing these deaths.

## Materials and Method

### Study Design and Setting

We conducted a retrospective cross-sectional study using the registry database of the Clinical Toxicology Department, the referral poisoning center in Khorshid hospital, Isfahan, Iran, affiliated with Isfahan University of Medical Sciences. This study was approved by the ethics committee of the Review Board of Isfahan University of Medical Sciences (IR.MUI.MED.REC.1403.284). We included patients who were hospitalized at Khorshid Hospital with acute poisoning between March 20, 2019, and March 20, 2023, and who died as a result of poisoning, as recorded in the registry system. Patients with incomplete data in the registration system were excluded.

### Data Extraction

In this study, data were extracted by reviewing the records of patients who were poisoned and subsequently died between March 20, 2019, to March 20, 2023, at the Isfahan Clinical Toxicology Department of Khorshid Hospital. The collected data were recorded using a standardized data collection form. The form included the following variables: gender, age, marital status, level of education, nationality, city of residence, cause of poisoning death, type of substance involved, method of poisoning, route of poisoning (gastrointestinal, cutaneous, inhalation, or injection), time interval from exposure to the toxic substance to hospital transfer, time interval from poisoning to death, length of hospitalization, history of substance abuse, history of suicide attempts, and history of addiction.

### Statistical analysis

All the data recorded in the data collection form were entered into SPSS software version 26 (IBM Corp., Armonk, N.Y., USA). The Shapiro-Wilk test was used to assess normality. Descriptive statistics included the mean and standard deviation (SD) for normally distributed data, the median and interquartile range (IQR) for non-normally distributed quantitative data, and frequency and percentage for qualitative data. Comparisons were performed using one-way ANOVA or Kruskal-Wallis tests for multiple groups and independent sample t-tests or Mann-Whitney U tests for two groups. A p-value of less than 0.05 was considered statistically significant.

## Results

Between 2019 and 2023, 306 patients died from acute poisoning at Khorshid Hospital. Most were male (249, 81.4%) and Iranian (302, 96.7%), with a median age of 42 years (IQR: 31–59). The majority had a middle school education. Median times from exposure to hospital transfer and from poisoning to death were 4 hours (IQR: 1–12) and 48 hours (IQR: 16–120), respectively, with a median hospitalization length of 48 hours (IQR: 12–120) (Table 1).

**Table 1.** Frequency and median values of demographic factors.

| Demographic factors | Frequency | Percentage |
| --- | --- | --- |
| Gender (M:F) | 249:57 | 81.4:18.6 |
| Age | 42 [31-59] | N/A |
| Education | Middle-school | N/A |
| Nationality (Iranian: non-Iranian) | 302:4 | 98.69:1.31 |
| City (Isfahan: Other) | 235: 71 | 76.80: 23.20 |
| Marital Status (M:S:D) | 221:84:1 | 72.22:27.45:0.33 |
| Method of poisoning (Suicide: Accidental: Abuse) | 193:58:55 | 63.07:18.95:17.97 |
| Route of exposure (Ingestion: Inhalation) | 289:17 | 94.44: 5.56 |
| Suicide history (N: Y) | 249:57 | 81.37: 18.63 |
| Abuse history (N: Y) | 276:30 | 90.20: 9.80 |
| Transfer (hours) | 4 [1-12] | N/A |
| Death time (hours) | 48 [16-120] | N/A |
| Hospitalization (hours) | 48 [12-120] | N/A |
| Addiction history |  |  |
| Amphetamine | 13 | 4.25 |
| Ethanol | 24 | 7.84 |
| Heroin | 14 | 4.57 |
| Methadone | 31 | 10.13 |
| Morphine | 1 | 0.33 |
| Opium | 50 | 16.34 |
| Cigarette smoking | 28 | 9.15 |
| None | 145 | 47.39 |

M: Male, F: Female, M: Married, S: Single, D: Divorce, N: No, Y: Yes, N/A: Not applicable

Suicide was the leading cause of poisoning (193 cases, 63.07%). The most frequently involved substances were methadone (63, 20.56%), aluminum phosphide (57, 18.63%), and paraquat (53, 17.32%) (Table 2). Ingestion was the predominant route of exposure (289, 94.44%), followed by inhalation (17, 5.56%). Cardiac arrest was the most common complication leading to death (263, 85.95%) (Table 3).

**Table 2.** Frequency of substance involved in fatal poisoning.

| Substance involved | Number | Percentage |
| --- | --- | --- |
| Methadone | 63 | 20.56% |
| Aluminum phosphide | 57 | 18.63% |
| Paraquat | 53 | 17.32% |
| Multi-drug | 36 | 11.76% |
| Methanol | 29 | 9.48% |
| Opium | 29 | 9.48% |
| Organophosphate | 12 | 3.92% |
| Amphetamine | 8 | 2.61% |
| Benzodiazepine | 7 | 2.29% |
| Beta-blocker | 3 | 0.98% |
| Selective serotonin reuptake inhibitor | 2 | 0.65% |
| Carbon monoxide | 2 | 0.65% |
| Acetaminophen | 1 | 0.33% |
| Arsenic | 1 | 0.33% |
| Acetylsalicylic acid | 1 | 0.33% |
| Tricyclic antidepressant | 1 | 0.33% |
| Valproate sodium | 1 | 0.33% |

**Table 3.** Frequency of fatal poisoning cause.

| Cause of fatal poisoning | Number | Percentage |
| --- | --- | --- |
| Cardiac arrest | 263 | 85.95% |
| Respiratory distress | 13 | 4.25% |
| Multi-organ failure | 6 | 1.96% |
| Sepsis | 5 | 1.63% |
| Brain death | 3 | 0.98% |
| End-stage renal disease | 2 | 0.65% |
| Pneumonia aspiration | 2 | 0.65% |
| Seizure | 2 | 0.65% |
| Anaphylaxis | 1 | 0.33% |
| Chronic obstructive pulmonary disease | 1 | 0.33% |
| Coronavirus disease | 1 | 0.33% |
| Gastrointestinal bleeding | 1 | 0.33% |
| Intracerebral hemorrhage | 1 | 0.33% |
| Mediastinitis | 1 | 0.33% |
| Naloxone withdrawal | 1 | 0.33% |
| Pulmonary Thromboembolism | 1 | 0.33% |
| Rhabdomyolysis | 1 | 0.33% |
| Subarachnoid hemorrhage | 1 | 0.33% |

A history of prior suicide attempts was reported in 57 patients (18.63%), while 30 patients (9.8%) had a history of substance abuse. Addiction history was present in 161 patients (52.61%), most commonly opium (50), methadone (31), and cigarette smoking (28).

Substance type was significantly associated with education level (P < 0.008) but not with gender or marital status. Route of exposure was associated with gender (P < 0.05) but not with marital status or education level. Substance abuse history was linked to gender (P < 0.03) and education (P < 0.05), while history of suicide attempts was associated with gender (P < 0.006). No significant associations were found between suicidal attempt history and marital status or education level. Also, no significant associations were observed between addiction history and gender, marital status, or education (Table 4 and Table 5).

**Table 4.**
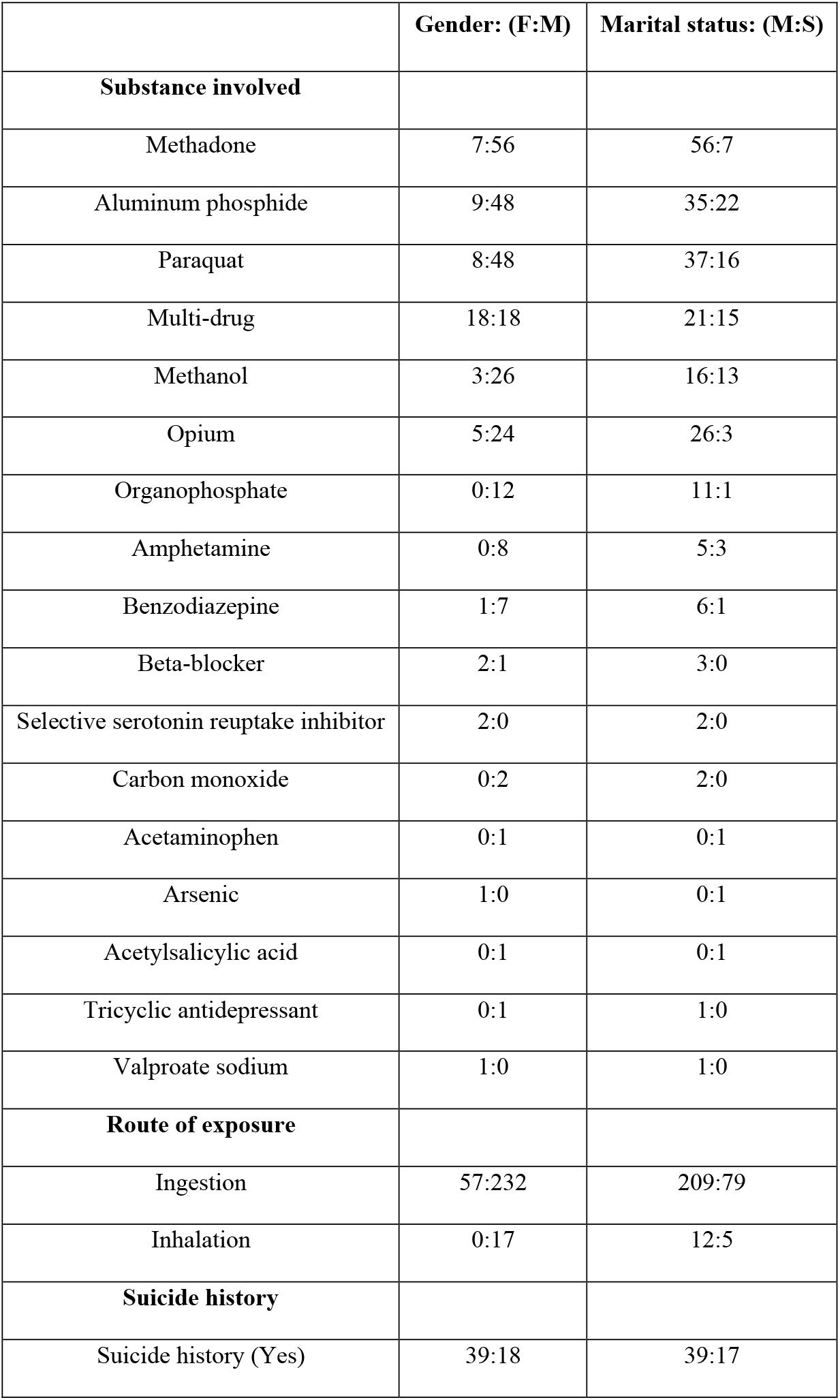
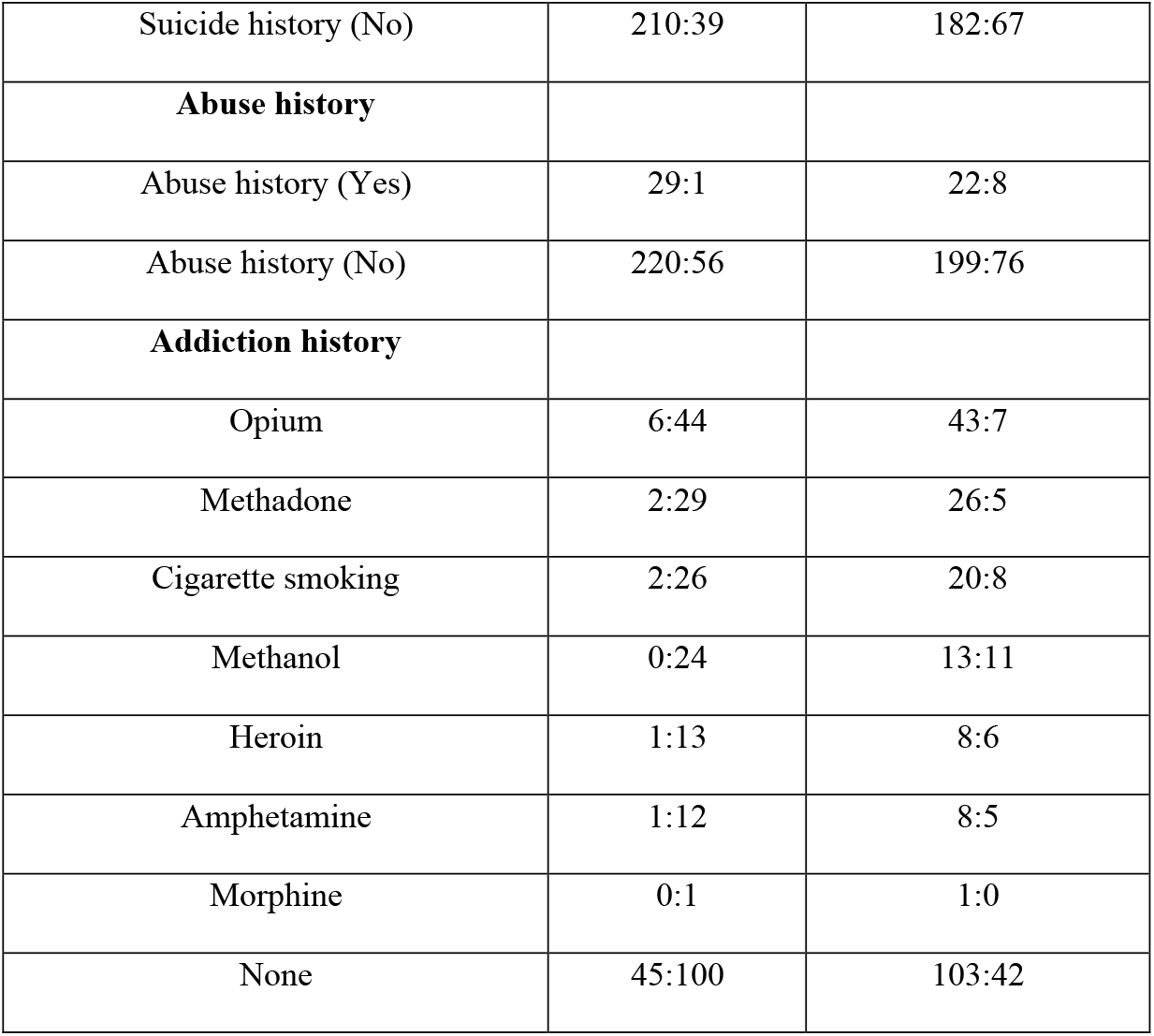
Distribution of poisoning-related variables by gender and marital status.

|  | **Gender: (F:M)** | **Marital status: (M:S)** |
| --- | --- | --- |
| **Substance involved** |  |  |
| Methadone | 7:56 | 56:7 |
| Aluminum phosphide | 9:48 | 35:22 |
| Paraquat | 8:48 | 37:16 |
| Multi-drug | 18:18 | 21:15 |
| Methanol | 3:26 | 16:13 |
| Opium | 5:24 | 26:3 |
| Organophosphate | 0:12 | 11:1 |
| Amphetamine | 0:8 | 5:3 |
| Benzodiazepine | 1:7 | 6:1 |
| Beta-blocker | 2:1 | 3:0 |
| Selective serotonin reuptake inhibitor | 2:0 | 2:0 |
| Carbon monoxide | 0:2 | 2:0 |
| Acetaminophen | 0:1 | 0:1 |
| Arsenic | 1:0 | 0:1 |
| Acetylsalicylic acid | 0:1 | 0:1 |
| Tricyclic antidepressant | 0:1 | 1:0 |
| Valproate sodium | 1:0 | 1:0 |
| **Route of exposure** |  |  |
| Ingestion | 57:232 | 209:79 |
| Inhalation | 0:17 | 12:5 |
| **Suicide history** |  |  |
| Suicide history (Yes) | 39:18 | 39:17 |
| Suicide history (No) | 210:39 | 182:67 |
| **Abuse history** |  |  |
| Abuse history (Yes) | 29:1 | 22:8 |
| Abuse history (No) | 220:56 | 199:76 |
| **Addiction history** |  |  |
| Opium | 6:44 | 43:7 |
| Methadone | 2:29 | 26:5 |
| Cigarette smoking | 2:26 | 20:8 |
| Methanol | 0:24 | 13:11 |
| Heroin | 1:13 | 8:6 |
| Amphetamine | 1:12 | 8:5 |
| Morphine | 0:1 | 1:0 |
| None | 45:100 | 103:42 |

**Table 5.** Associations between demographic factors with substance type, route of poisoning, and behavioral histories.

|  | Gender | Marital-status | Education-level |
| --- | --- | --- | --- |
| Type of substance | $P = 0.25$<br>$U = 7778.50$ | $P = 0.16$<br>$Test-statistic = 3.67$ | $P < 0.008^*$<br>$Test-statistic = 15.892978$ |
| Route of poisoning | $P < 0.05^*$<br>$U = 6612.00$ | $P = 0.96$<br>$Test-statistic = 0.09$ | $P = 0.66$<br>$Test-statistic = 3.26$ |
| Abuse history | $P < 0.03^*$<br>$U = 6394.50$ | $P = 0.94$<br>$Test-statistic = 0.12$ | $P < 0.05^*$<br>$Test-statistic = 11.22$ |
| Suicide history | $P < 0.006^*$<br>$U = 5967.00$ | $P = 0.10$<br>$Test-statistic = 4.63$ | $P = 0.65$<br>$Test-statistic = 3.33$ |
| Addiction history | $P = 0.53$<br>$U = 7449.00$ | $P = 0.08$<br>$Test-statistic = 4.96$ | $P = 0.29$<br>$Test-statistic = 6.19$ |

\* $P < 0.05$

## Discussion

This study identified 306 poisoning-related deaths, with suicide being the leading cause. The most commonly involved substances were methadone, aluminum phosphide, and paraquat, with ingestion as the primary route of exposure. Analysis of demographic factors revealed that male gender, middle age, lower education level, and a history of addiction were significantly associated with fatalities. Cardiac arrest was the most common complication leading to death.

### Global Patterns of Poisoning Mortality

The poisoning mortality patterns observed in this study align with findings from previous research. Globally, suicide remains a leading cause of fatal poisoning. A study in north-eastern India reported that 63.96% of 505 poisoning-related deaths were due to suicide (Sharma et al., 2019).

Similarly, an analysis of poisoning deaths in eastern Iran between 2010 and 2016 found that suicide accounted for 38.2% of cases (Mehrpour et al., 2018). In Isfahan, Iran, a prior study (2014–2019) reported that 63.2% of poisoning-related deaths were suicide, a finding consistent with the present study (Dorooshi et al., 2022).

However, patterns of poisoning-related mortality vary across regions. In the United States, an analysis of poisoning deaths from 2006 to 2015 found that 84.7% of 36,422 cases were unintentional (Ali et al., 2019). This suggests that while suicide is a major cause of poisoning-related deaths worldwide, regional differences exist due to variations in substance availability, mental health factors, and healthcare accessibility.

### Common Substances Involved in Fatal Poisoning

The substances responsible for poisoning fatalities vary across different regions. In this study, methadone was the most commonly involved substance. In contrast, a cross-sectional study in Ireland (2004–2017) identified cocaine as the leading cause of fatal poisoning (Lynn et al., 2021). A study in north-eastern India found that organophosphates were the primary cause of poisoning-related deaths (Sharma et al., 2019), while research in Japan identified paraquat and organophosphates as the major toxic agents (Lee et al., 2008).

In Iran, aluminum phosphide has been a significant contributor to poisoning deaths. A study conducted in Mazandaran (2011–2016) reported that 65.5% of poisoning-related deaths were due to aluminum phosphide (Rezaei et al., 2020). Similarly, research conducted at Baharloo Hospital in Tehran (2011–2014) identified aluminum phosphide as the leading cause of poisoning fatalities (Titidezh et al., 2019). A previous study at Khorshid Hospital in Isfahan (2014–2019) found that paraquat and aluminum phosphide were the most frequently involved substances (Dorooshi et al., 2022). However, in our study, while aluminum phosphide remained a major contributor, ranking as the second most common substance involved in fatal poisonings, methadone was the leading cause. The prominent role of methadone is likely related to its long half-life and widespread availability. These findings highlight the need for careful monitoring and potentially longer hospitalization for patients with methadone poisoning.

### Demographic Factors Associated with Fatal Poisoning

Demographic factors related to poisoning mortality vary across studies. Although poisoning incidence is often higher in females, mortality rates are generally higher in males, likely due to higher-risk behaviors such as substance abuse. Fatal poisonings occur across all age groups, but adults are more frequently affected than younger or older populations. A study in the United States (2003–2007) found that among both unintentional and suicidal poisoning deaths, males, Caucasians, and individuals aged 40–59 had the highest mortality rates (Muazzam et al., 2012). Similarly, research in British Columbia (2008–2013) reported 3,120 poisoning-related deaths, with 66% of fatalities occurring in males, while 59% of hospitalizations involved females (Pawer et al., 2021).

In Ilam, Iran (1993–2013), poisoning-related suicide attempts were more common among females, single individuals, those with lower education, and those aged 15–24 years, whereas completed suicides were more frequent among males (Azizpour et al., 2016). Our study similarly found higher mortality among males and individuals with lower education levels, while married individuals and those aged 31–55 were most affected. A previous study at Khorshid Hospital in Isfahan (2014–2019) reported that 73.1% of poisoning fatalities occurred in males, with an average age range of 22–63 years, consistent with our findings (Dorooshi et al., 2022). Additionally, that study, like ours, found a higher fatal poisoning rate among married individuals.

### Hospitalization Duration and Timing of Death

The length of hospitalization varies depending on the type of substance, the time from exposure to hospital admission, and other clinical factors. A retrospective study in Qatar reported a mean hospitalization duration of 1.84 days (Salem et al., 2024), which aligns with the median hospitalization time of two days in our study. In contrast, a study at Baharloo Hospital in Tehran (2011–2014) reported a hospitalization duration of 3–6 days, with most deaths occurring within 10 days of admission (Titidezh et al., 2019).

### Limitations and Future Directions

Despite the strengths of this study, including a comprehensive dataset, a few limitations should be addressed. The retrospective nature of data collection may introduce selection bias, and the absence of detailed psychiatric histories limits insights into underlying mental health conditions contributing to poisoning fatalities. Additionally, since the study was conducted in a single clinical center, the findings may not be fully generalizable to other regions. Future research should focus on multi-center studies to provide a broader perspective on poisoning trends and their socio-economic determinants. Investigations into psychiatric comorbidities, access to toxic substances, and the effectiveness of suicide prevention programs are also needed. Moreover, prospective studies evaluating early interventions, emergency response efficiency, and novel treatment protocols could provide valuable insights into reducing poisoning-related deaths. Strengthening policies to regulate highly toxic substances, particularly methadone and aluminum phosphide, should be a priority in public health strategies.

## Conclusion

This study highlights the significant burden of poisoning-related mortality, with suicide as the leading cause and methadone as the most frequently involved substance. The findings highlight the need for stricter regulations on methadone distribution, improved mental health interventions, and strengthened substance abuse prevention programs. Given the high mortality associated with aluminum phosphide and paraquat, preventive measures, including tighter control over pesticide availability, should be considered.

Considering the demographic patterns observed, targeted awareness movements and early intervention strategies should be prioritized for high-risk populations. By addressing these challenges, the overall impact of poisoning-related deaths and public health outcomes could be improved.

## Data availability

The data will be available upon reasonable request from corresponding author.

## Ethics declaration

This study was approved by the ethics committee of the Review Board of Isfahan University of Medical Sciences (IR.MUI.MED.REC.1403.284)

## Acknowledgment

N/A.

## Authors contribution

Conceptualization: N.Y, G.D, N.E. Dataset preparation: N.Y, G.D, N.E. Statistical analysis: N.Y, G.D, N,E. Data interpretation: N.Y, S.S, G.D, N.E. Drafting original manuscript: N.Y. Revising the manuscript: S.S, G.D, N,E. All the authors have read and approved the final version for publication and agreed to be responsible for the integrity of the study.

